# Comparative Effectiveness of ACE Inhibitors and Angiotensin Receptor Blockers for Cardiovascular Disease Prevention in People Living with HIV and Hypertension

**DOI:** 10.64898/2026.01.12.26343966

**Authors:** Tom Forzy, Henock G. Yebyo, Gregory M. Lucas, Huldrych F. Günthard, Catherine R. Lesko, Vincent C. Marconi, Timothy R. Sterling, Michael J. Silverberg, Maile Y. Karris, Michael A. Horberg, Sonia Napravnik, Keri N. Althoff, Milo A. Puhan

**Affiliations:** Epidemiology, Biostatistics and Prevention Institute, Department of Epidemiology, University of Zurich, Zurich, Switzerland; Johns Hopkins University School of Medicine, Baltimore, Maryland, USA; Department of Infectious Diseases and Hospital Epidemiology, University Hospital Zurich, Zurich, Switzerland; Institute of Medical Virology, University of Zurich, Zurich, Switzerland; Johns Hopkins Bloomberg School of Public Health, Department of Epidemiology, Baltimore, Maryland, USA; Division of Infectious Diseases, Emory University School of Medicine and Department of Global Health, Rollins School of Public Health, Atlanta, GA, USA; Division of Infectious Diseases, Vanderbilt University Medical Center, Nashville, TN; Division of Research, Kaiser Permanente Northern California, Pleasanton, CA; Division of Infectious Diseases and Global Public Health, University of California San Diego, San Diego, California, USA; Kaiser Permanente and Care Management Institute, Mid-Atlantic Permanente Medical Group, Washington DC, USA; University of North Carolina at Chapel Hill, Chapel Hill, North Carolina, United States of America

**Keywords:** Anti-hypertensive drugs, Angiotensin-converting enzyme inhibitors, Angiotensin receptor blockers, Cardiovascular diseases, HIV, Trial emulation

## Abstract

**Background:** Angiotensin-converting enzyme inhibitors (ACEIs) and angiotensin receptor blockers (ARBs) are established antihypertensive treatments that reduce cardiovascular disease (CVD) risk. However, their comparative effectiveness in people with HIV (PWH) is not well examined. This study evaluated the comparative effectiveness of ACEIs and ARBs head-to-head and versus no antihypertensive treatment in preventing primary CVD.

**Methods:** Using a target trial emulation framework and data from the North American AIDS Cohort Collaboration on Research and Design (NA-ACCORD), we estimated observational analogs of intention-to-treat (ITT) and per-protocol (PP) effects of antihypertensive treatments in preventing primary CVD (myocardial infarction, non-MI coronary artery disease, stroke, transient ischemic attack, peripheral vascular disease, cardiovascular death) among hypertensive PWH, with subgroup analyses for Black and White PWH.

**Results:** Compared with no antihypertensive treatment, ACEIs and ARBs were both associated with lower CVD risk in PWH, with similar effect sizes in ITT and PP analyses (ACEI ITT adjusted hazard ratio (HR): 0.79, 95% CI [0.70-0.89]; ACEI PP: 0.71 [0.55-0.90]; ARB ITT: 0.87 [0.65-1.16]; ARB PP: 0.37 [0.18-0.76]). Race-stratified ITT and PP analyses suggested somewhat greater risk reductions in White than Black PWH, although differences were not statistically significant. In head-to-head comparisons, ACEIs and ARBs showed comparable effectiveness overall (ITT: 1.14 [0.84-1.55]; PP: 0.54 [0.25-1.18]), and within race strata.

**Conclusions:** Our study found that both ACEIs and ARBs were effective in reducing CVD risk among PWH, with similar effectiveness observed for both medications. The analysis did not reveal statistically significant differences in effectiveness between Black and White PWH for either drug.

## 1. Introduction

The role of anti-hypertensive drugs, and specifically angiotensin-converting enzyme inhibitors (ACEIs) and angiotensin receptor blockers (ARB), in the prevention of cardiovascular diseases (CVD) in the general population, has been a subject of extensive research over the past decades (1–7), including randomized trials. These classes of drugs reduce the risk of major CVD events, including myocardial infarction (MI). Both ACEIs and ARB reduce mortality and morbidity in placebo-controlled trials (8–11). Although not well examined, a recent study in the general population showed no difference in effectiveness for CVD prevention between the two drug classes (12), but ARB users experienced less adverse effects than ACEI users (5,6,8).

Studies on CVD prevention for high-risk populations are scarce, particularly for people living with HIV (PWH). PWH have twice as much risk of CVD as compared with the general population, stimulated by chronic inflammation and some antiretroviral drugs (e.g. abacavir), in addition to the traditional risk factors (13). Although statins are recommended for PWH with moderate or high CVD risk, statins do not lower blood pressure (14). The potential impact of ACEIs and ARBs for CVD prevention in this group is not sufficiently explored (15). A recent study in a cohort of US Veterans with HIV and hypertension highlighted the cardiovascular benefits of ACEIs and ARBs over other anti-hypertensive drugs in PWH (16). But to our knowledge, no study has directly compared the effectiveness of ACEIs versus ARBs for CVD prevention in PWH.

Additionally, race has been identified as a potential effect modifier in the efficacy of ACEIs and ARBs in many studies among people without HIV, with evidence suggesting that these drugs may have reduced effects in Black individuals (17–20). Weir et al. suggest that this differential response could be partly explained by physiological differences, with hypertensive patients of African descent being more likely to have a low-renin, sodium-sensitive profile and greater plasma volume expansion, in contrast to White patients, who more commonly have higher renin levels and may respond better to ACEIs (21,22). J.A. Johnson also raises the potential role of pharmacogenetics (23). While studies have not yet identified consistent genetic variants that determine this response, it is hypothesized that genotype frequencies affecting drug responsiveness may differ among ancestries (23). Moreover, sociocultural factors such as diet, can further influence antihypertensive drug efficacy (23). Despite Black individuals being disproportionately represented among PWH, no studies have specifically examined racial differences in the effectiveness of ACEIs and ARBs among PWH.

In this study, we used a target trial emulation framework to estimate the comparative effectiveness of ACEIs and ARBs individually for primary CVD prevention, using North American AIDS Cohort Collaboration on Research and Design (NA-ACCORD) data. Additionally, we evaluated and compared the effects of both drugs against a third group with diagnosed hypertension but not receiving any anti-antihypertensive treatment, and we investigated potential differences in the effectiveness of ACEIs and ARBs in Black PWH compared to White PWH.

## 2. Methods

### 2.1 Study design

We first developed a hypothetical open-label target trial protocol for estimating the effectiveness of ACEIs versus ARBs for primary prevention of CVDs in PWH (Table 1). We emulated the target trial protocol, including the eligibility, treatment assignment, and outcomes using data from the clinical cohorts of the NA-ACCORD.

**Table 1.** Specification and emulation of an open label target trial of ACEIs versus ARBs on the risk of first CVD event in people with HIV.

| Specification and Protocol component | Target trial specification | Target trial emulation |
| --- | --- | --- |
| Eligibility criteria | Having HIV and diagnosed hypertension | Same as for the target trial |
|  | No history of any antihypertensive prescription or major CVD event |  |
|  | Define baseline as the first month in which eligibility criteria are met |  |
| Treatment strategies | Initiation of first-line anti-hypertensive treatment with either (1) an ACEI or (2) an ARB, as well as continuation of the treatments over follow-up for the per-protocol analysis | Same as for the target trial. We considered treatment to be discontinued if there was a gap of more than 1 month between successive prescriptions |
| Treatment assignment | Individuals are randomly assigned to a strategy at baseline and will be aware of the strategy to which they have been assigned | We classified individuals according to the strategy that their data were compatible with at baseline and attempted to emulate randomization by adjusting for baseline confounders |
| Outcomes | First CVD event | Same as for the target trial |
| Follow-up | Starts at baseline and ends at the month of first CVD event diagnosis, death, administrative censoring, or 5 years of follow-up, whichever happens first. Per-protocol analysis: censor participants if and when they deviate from their assigned treatment strategy | Same as for the target trial |
| Causal contrasts | Per-protocol and intention-to-treat effects | Observational analog of intention-to-treat and per-protocol effects |
| Statistical analysis | Per-protocol and intention-to-treat analysis.<br>Apply inverse-probability weights to adjust for factors associated with adherence.<br>Pooled logistic regression to estimate intention-to-treat and per protocol effects via hazard ratios and standardized survival curves | Same per-protocol and intention-to-treat analyses with emulation and additional adjustment for baseline covariates |

### 2.2 Data

Established as part of the International Epidemiologic Databases to Evaluate AIDS (IeDEA) initiative, the NA-ACCORD data allows for investigations that require large sample sizes and heterogeneity among the study population. The NA-ACCORD pools individual-level data from >20 longitudinal cohorts of PWH from the United States and Canada (24). The contributing cohorts are geographically and demographically diverse and include academic and community-based clinical cohorts. PWH in contributing clinical cohorts who successfully link into HIV care, defined as ≥2 HIV clinical visits within 12 months, are eligible to be enrolled in the NA-ACCORD. NA-ACCORD participants provided informed consent or contributed data with a waiver of informed consent with approval of local institutional review boards. The demographics of PWH in the NA-ACCORD are reflective of people with diagnosed HIV infection in the United States (25).

NA-ACCORD collects data elements from participating cohorts via an annual standardized protocol, including demographic characteristics, HIV risk factors, clinical diagnoses, laboratory measurements (such as HDL and triglycerides), medication histories (including antiretroviral therapy and medications for chronic diseases), vital status (through clinical tracking and linkages to local, provincial and national death registries), and healthcare utilization. Data are submitted to the Data Management Core for harmonization and quality assurance and then transferred to the Epidemiology/Biostatistics Core where analytic-ready variables are created (24). The protection of human subjects in the NA-ACCORD is overseen by local institutional review boards and at the Johns Hopkins Bloomberg School of Public Health.

We included PWH who were alive and under follow-up (using an observation window approach) during our study period from January 1, 1995 and December 31, 2023. Observation windows are defined by the Epidemiology and Biostatistics Core of the NA-ACCORD during data harmonization as periods during which NA-ACCORD cohorts provided complete data on relevant covariates—in this case, diabetes diagnoses and medications, hypertension diagnoses and medications, lipid measurements and medications, and BMI.

### 2.3 Eligibility criteria for our nested study

In this target trial emulation, we restricted our analysis to PWH with a diagnosis of hypertension, observed covariate values, and no prior reported history of prescription antihypertensive medication (see Web Appendix). Eligibility to the treatment group (and baseline of follow-up) was defined as the first month in which an individual initiated an ACEI or ARB within this observation window. Eligibility to the control group (and baseline of follow-up) was defined at the first observed month following a diagnosis of hypertension, within this observation window and without any history of antihypertensive therapy or previous diagnoses of major CVD (Myocardial Infarction (MI), non-MI coronary artery disease, stroke, transient ischemic attack (TIA), peripheral vascular disease, and cardiovascular death). This definition of eligibility allowed for up to two eligibility months per individuals, leading to the successive enrollment in first the control group and then the treatment group for all individuals who had first a diagnosed hypertension without antihypertensive treatment initiation, and who later initiated ACEI or ARB as a first antihypertensive treatment.

### 2.4 Treatment assignment

We compared three hypothetical dynamic treatment strategies and estimated both intention-to-treat (ITT) and per-protocol (PP) effects. For the ITT analysis, we compared (i) the initiation at baseline of only ACEIs versus the initiation of only ARBs, (ii) the initiation of only ACEIs at baseline versus no-initiation of any anti-hypertensive treatment at baseline, and (iii) the initiation of only ARBs at baseline versus no-initiation of any anti-hypertensive treatment at baseline.

For the PP analysis, we compared (i) the initiation of only ACEIs at baseline and continuation over follow-up versus the initiation of only ARBs at baseline and continuation over follow-up, (ii) the initiation and continuation of only ACEIs versus no-initiation of any anti-hypertensive treatment at any time over follow-up, and (iii) finally the initiation of only ARBs versus no-initiation of any anti-hypertensive treatment at any time over follow-up. In the PP analysis, we right-censored patients when they deviated from their assigned treatment strategy. In the groups initiating ACEIs and ARBs, patients were right censored upon discontinuation of the assigned medication (defined as a gap of ≥1 month in prescribed medication) or initiation of a different class of antihypertensive therapy during follow-up. In the group not initiating anti-hypertensive treatment, we right-censored patients when they initiated any anti-hypertensive medication during follow-up.

Conditional exchangeability between groups was assumed, with adjustments made both for various baseline (ITT) and time-varying (PP) demographic and clinical confounding factors: age, sex, BMI, smoking status, risk for HIV acquisition and transmission (men having sex with men, injection drug use), laboratory measurements (HDL, triglycerides), diagnoses (hepatitis C, chronic kidney disease (CKD), diabetes), and medications (specific antiretroviral treatment (ART), anticoagulants and platelet agents, and statins). For CKD, we used documented diagnosis, and for diabetes, we applied the NA-ACCORD operational definition, classifying patients as diabetic at and after an HbA1c >6.5%, or a prescription for a diabetes-specific medication, or a documented diabetes diagnosis and a prescription for a diabetes-related medication.

### 2.5 Outcomes

The primary outcome of CVD event was defined as the first observed occurrence of MI, non-MI coronary artery disease, stroke, transient ischemic attack (TIA), peripheral vascular disease, and cardiovascular death (i.e., cardiovascular event as the underlying cause of death). The procedure for validating MIs within NA-ACCORD has been described in detail (26). However, for the other CVD event types, events were ascertained from diagnosis codes and were not further validated. (24)

### 2.6 Follow-up

Patients were followed up from the first month in which they met the eligibility criteria until their study exit, defined as the first occurrence of any these events: [1] outcome of interest; [2] death from another cause; [3] administrative censoring; [4] closing of cohorts or observation window for hypertension medication, diabetes medication, BMI, lipids medication, and CVD (see Webappendix for detailed cohort observation windows (27)); [5] completion of 5-years follow-up; [6] loss to clinical follow-up, defined as occurring after 24 months with no clinical visits, viral load measurement or CD4 measurement; and [7] in the PP analysis, patients were right-censored at the time they deviated from their baseline treatment strategy.

### 2.7 Statistical analysis

We employed a discrete time-to-event analysis at the month level. To control for confounding, our outcome model included adjustment for baseline covariates. Specifically, we included all potential confounders of the effect of treatment assignment on the outcome, measured at the start of the emulated trial for each individual (28). To address potential selection bias due to death, administrative censoring, or loss to follow-up, we additionally applied inverse probability of censoring weighting (IPCW). For the PP analysis, we further adjusted for selection bias arising from censoring at per-protocol violations by applying a second set of IPCWs. Both sets of inverse probability weights were estimated using pooled logistic regression models for the probability of being censored, and weights were constructed as the cumulative product of month-specific covariate-conditional probabilities of remaining uncensored. For the ITT analysis, the final weights consisted of the IPCW for censoring. For the PP analysis, the final weights were the product of the censoring IPCW and the IPCW for per-protocol adherence. To reduce the impact of extreme weights, all weights were truncated at the 99th percentile (28). We checked that the mean of the IPW was approximately one as a quality control to ensure proper standardization and interpretation of the weighted sample.

Finally, we fitted weighted pooled logistic regression models for the effects of (a) ACEIs versus ARBs; (b) ACEIs versus no antihypertensive treatment; and (c) ARBs versus no antihypertensive treatment. Because there was a low monthly hazard of CVD events, the odds ratio from this model approximated the hazard ratio (HR) (29). We estimated 95% confidence intervals (CI) for the HRs using a robust sandwich estimator.

We estimated marginal treatment effects by fitting three weighted pooled logistic regression models for the three emulated trials. Using each fitted model, we predicted the cumulative incidence at each time point for every individual under two hypothetical scenarios: (1) assuming assignment to the treatment group, and (2) assuming assignment to the control group (30). For each scenario, we then calculated the average cumulative incidence across all individuals. The 5-y marginal risk difference (RD) was defined as the difference between the average predicted cumulative incidences under the two scenarios (i.e., average cumulative incidence for the treated group minus that for the control group). Finally, we derived simulation-based confidence intervals (CIs) by sampling 500 times from the asymptotic distribution of the model coefficients (31).

Subgroup analyses were conducted for Black and White patients. We conducted both ITT and PP subgroup analysis for ACEI initiators versus no treatment, but only an ITT analysis for ARB initiators versus no treatment and ARB versus ACEI initiators due to the limited number of ARB initiators (see Table 2). To test our hypothesis of differential effectiveness by race, we restricted to Black and White individuals, encoded race as a dummy variable, and reported the p-value of the coefficient on the product term between treatment and race in the pooled logistic regression model.

**Table 2.**
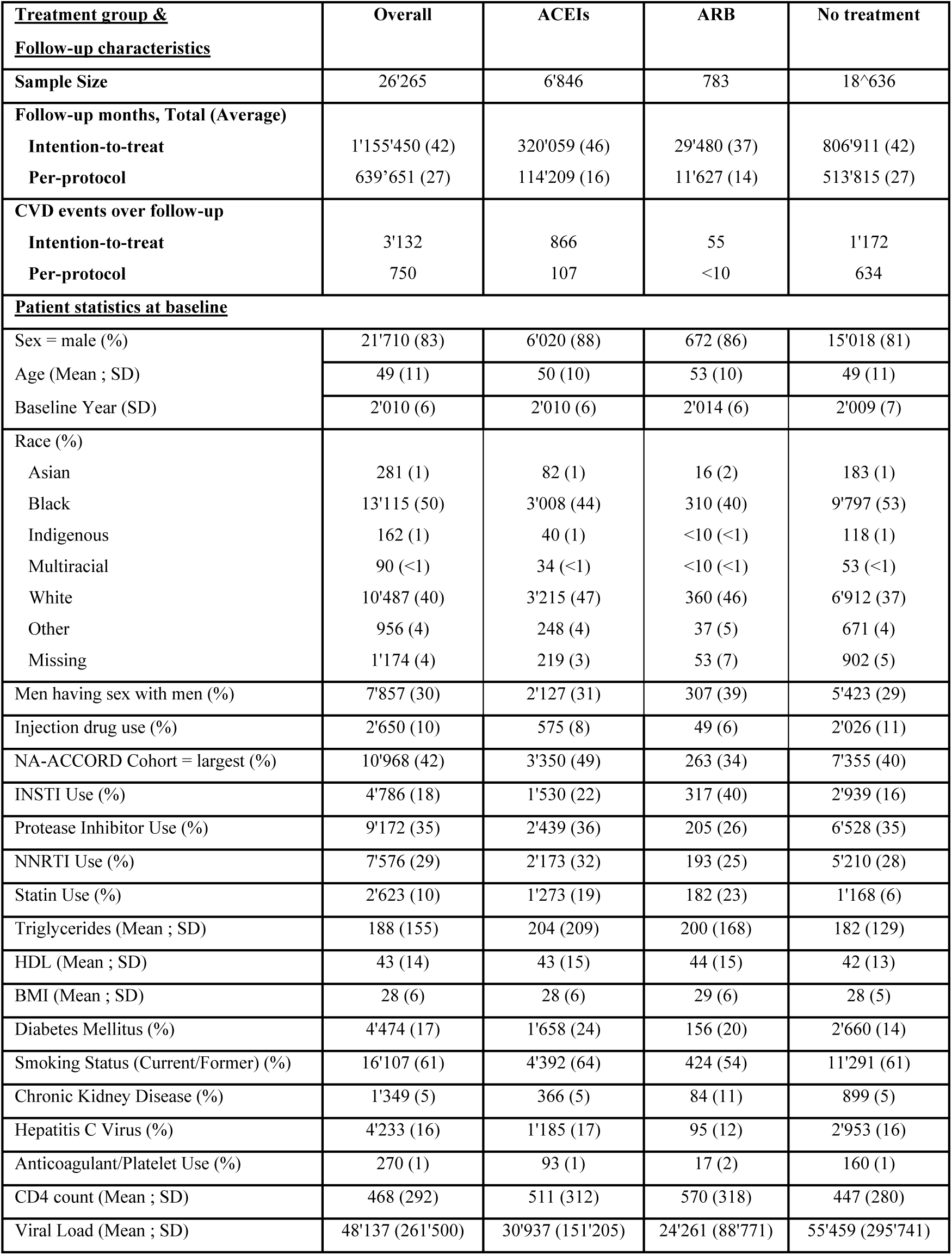
Cohort, follow-up and patient characteristics at baseline of the emulated trials comparing three groups: hypertensive individuals initiating ACEIs, initiating ARBs or not initiating any anti-hypertensive treatment.

All analyses were conducted using R software version 4.5.0 (www.r-project.org).

### 2.8 Sensitivity analysis

To test the robustness of our findings and evaluate potential cohort-specific effects, we conducted a sensitivity analysis excluding the largest contributing cohort to ensure their contribution was not masking the relationships of our exposures and outcomes in the other contributing cohorts due to differences in sociodemographic and clinical practice. We repeated all primary and secondary analyses on this restricted sample.

## 3. Results

Out of a total of 165,995 PWH included in the diagnosis data of NA-ACCORD between 1995 and 2024, 63,875 were diagnosed with hypertension. We then identified individuals eligible for our emulated trials according to the criteria listed in Table 1. Of these individuals, 6,846 initiated ACEIs. The mean follow-up time under the ITT analysis (measuring follow-up with HIV care and within observation windows and allowing for treatment deviation) was 46 months, and the median was 60 months, meaning that at least half of the individuals in this group had a complete follow-up). The mean follow-up was 16 months under the PP analysis (measuring follow-up while on ACEIs without allowing for treatment deviation, median: 7 months). Similarly, 783 individuals initiated ARBs, with mean follow-up of 37 months (median: 20) and 14 (median: 7) months for ITT and PP analyses, respectively. 18,636 individuals were included in the no-treatment group (no initiation of any antihypertensive medication at baseline); their mean follow-up was 42 months (ITT, median: 60) and 27 months (PP, median: 20) (see Table 2 for group characteristics). The large difference between ITT and PP duration of follow-up suggests that a large proportion of the individuals without a treatment assigned at baseline initiated anti-hypertensive treatment shortly after, and that individuals assigned to a treatment at baseline often initiated or switched to another treatment soon after. Those individuals were then right censored in the PP analysis.

Critical for interpretation of results, we note that although Black people made up 50% of the full sample, they accounted for only 4% of those treated with ACEIs and 40% of those treated with ARB. In contrast, White people made up 40% of the full sample, but 47% of people who initiated an ACEI and 46% of people who initiated an ARB. We also note that female patients were less likely to initiate an ACEI (12%) or ARB (14%) and more likely to be untreated (19%). ARB initiators were far more likely to be prescribed an INSTI-based ART regimen (40%) versus compared to ACEI initiators (22%) or untreated people (16%). Table 2 provides further detail on the sociodemographic and clinical profiles of each treatment arm at baseline, including the number of cases.

### ACEIs vs. no antihypertensive treatment

Results are presented in Figures 2,3 and Table 3. When compared to the absence of any antihypertensive treatment in the full sample of hypertensive PWH, ACEIs reduced the hazard of CVD by 21% in the ITT analysis (0.79 [0.70-0.89]), and by 29% in the PP analysis (0.71 [0.55-0.90]) (Figure 2, Table 2). 5-year RDs are given in Table 3.

**Figure 1.**
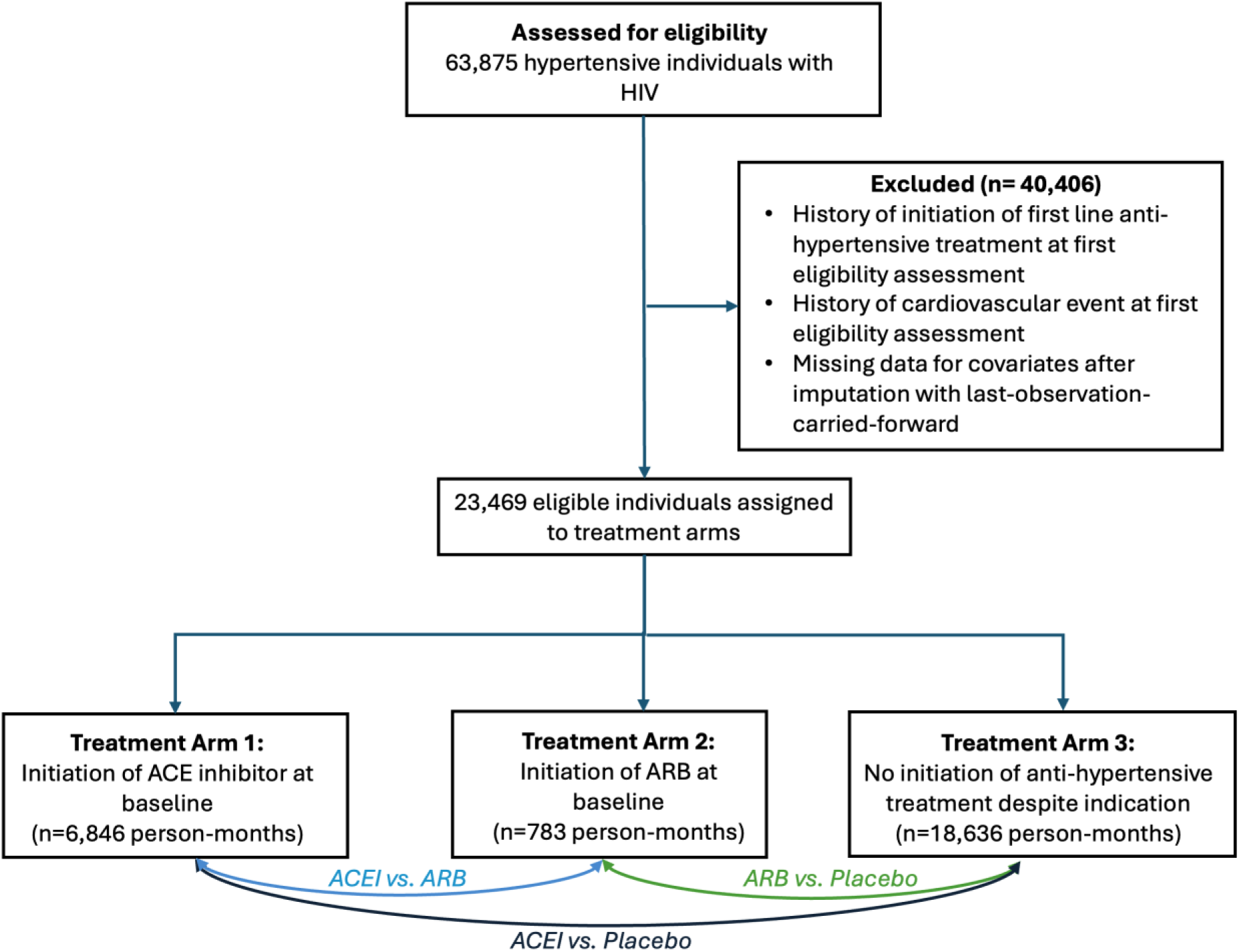
Flowchart illustrating the selection of eligible individuals from NA-ACCORD to emulate three target trials comparing CVD risk between two treatment groups : initiation of ACEI (Arm 1) versus initiation of ARB (Arm 2) ; initiation of ACEI (Arm 1) versus no-initiation of anti-hypertensive treatment at baseline (Arm 3), and initiation of ARB (Arm 2) versus no-initiation of anti-hypertensive treatment at baseline (Arm 3). Numbers in parentheses indicate unique individuals in each category.

**Figure 2.**
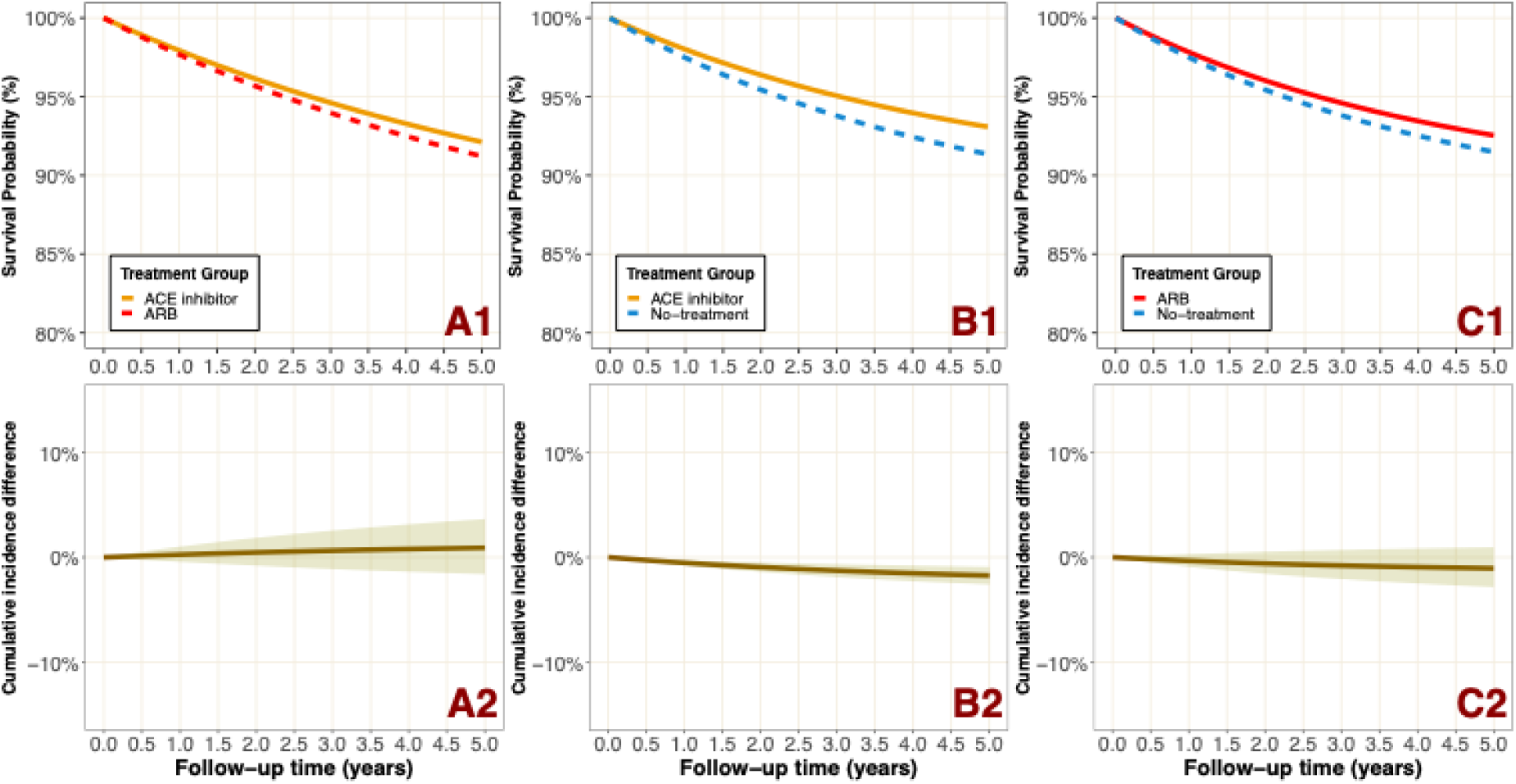
Standardized survival curves without cardio-vascular disease and difference in cumulative incidence for hypertensive people with HIV estimated from an intention-to-treat analysis between: (A1, A2) ARB versus ACEI ; (B1, B2) ACEI versus no antihypertensive treatment; (C1, C2) ARB versus no antihypertensive treatment. Based on NA-ACCORD data.

**Figure 3.**
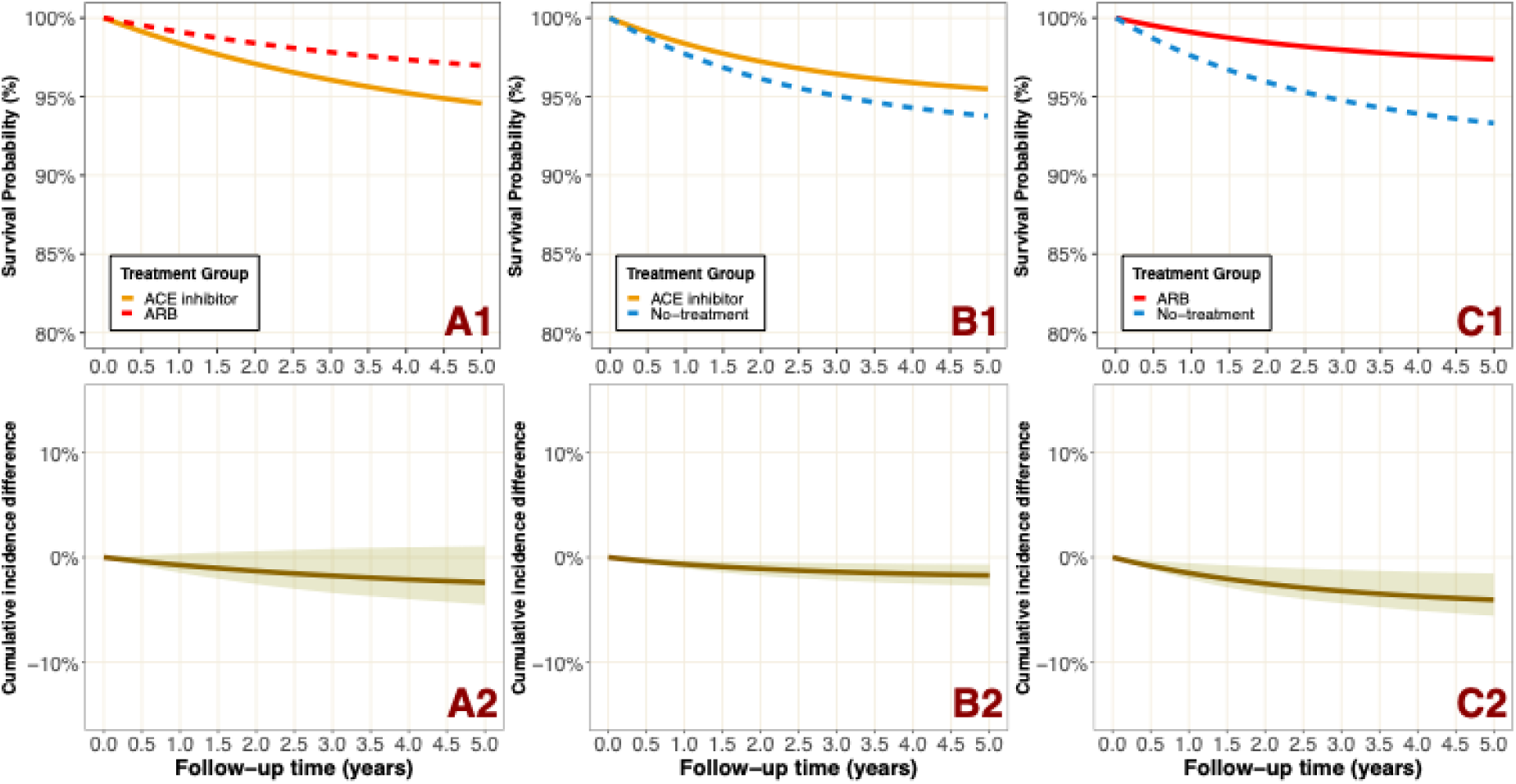
Standardized survival curves without cardio-vascular disease and difference in cumulative incidence for hypertensive people with HIV estimated from a per-protocol analysis between: (A1, A2) ARB versus ACEI ; (B1, B2) ACEI versus no antihypertensive treatment; (C1, C2) ARB versus no antihypertensive treatment. Based on NA-ACCORD data.

**Figure 4.**
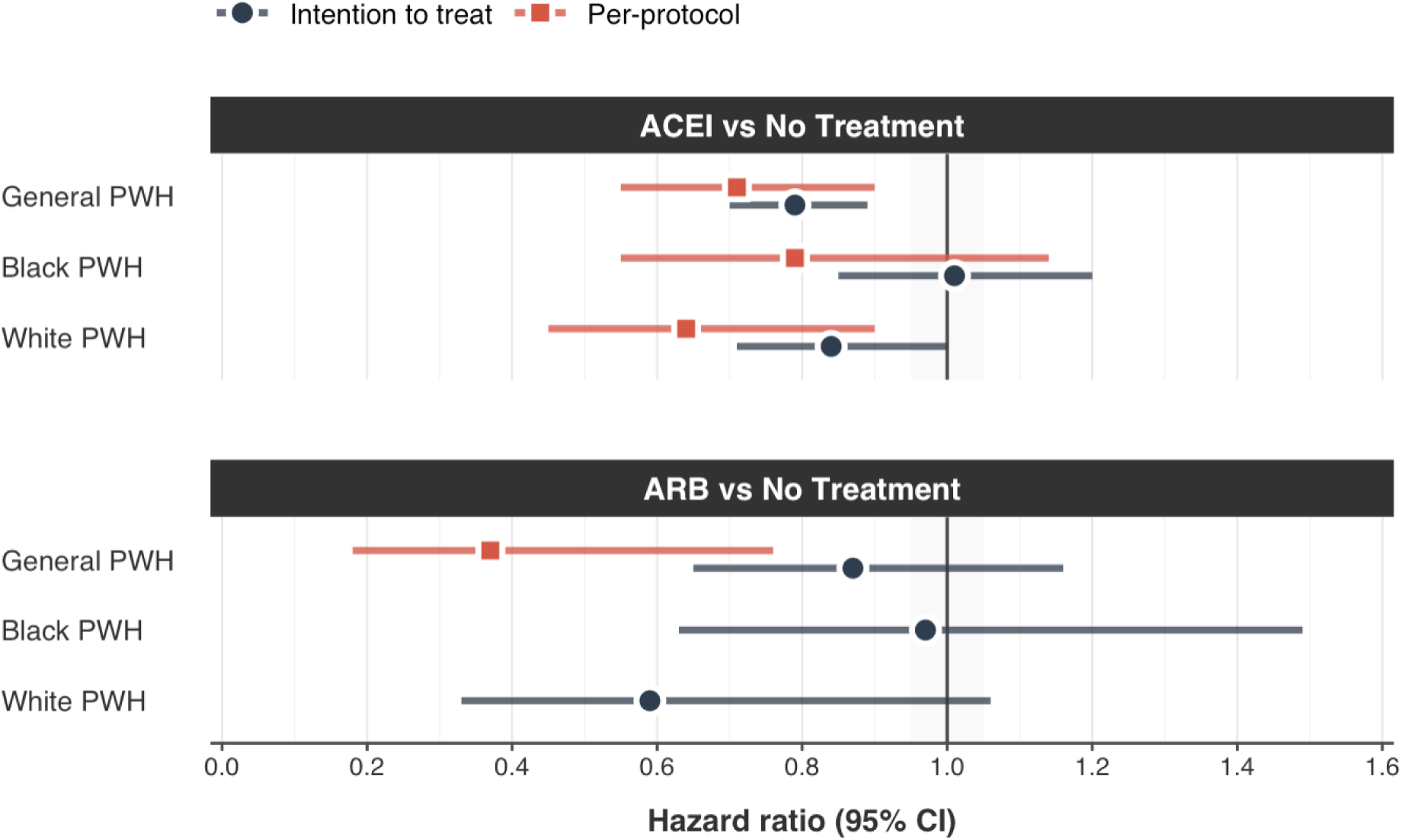
Forest plot of estimated hazard ratio of primary cardiovascular events obtained when emulating two target trials of angiotensin converting enzyme inhibitor (ACEI) versus no antihypertensive treatment, and angiotensin receptor blocker (ARB) versus no antihypertensive treatment on the risk of in people with HIV (PWH) and stratified by ethnicity.

**Table 3.**
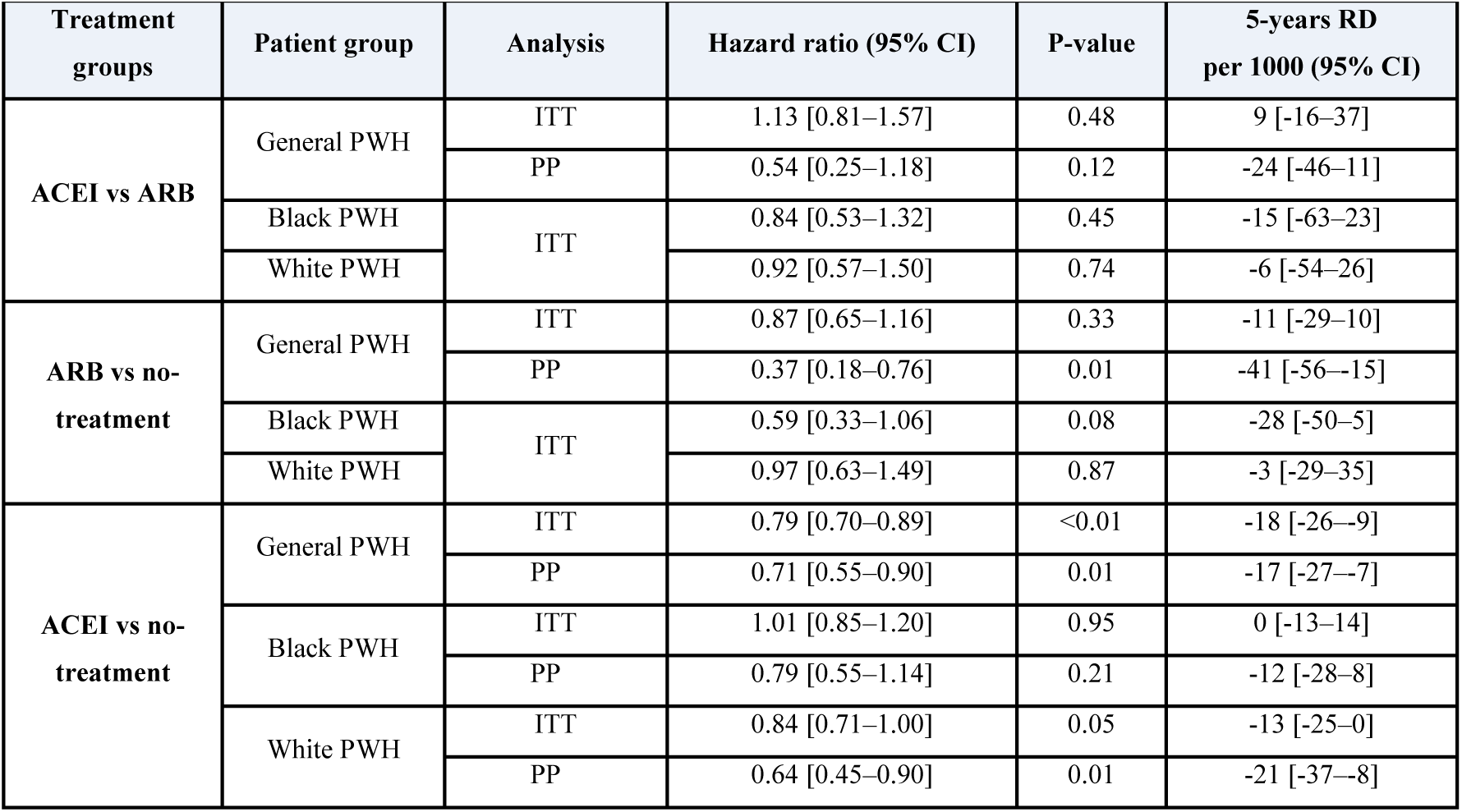
Estimated hazard ratio and risk differences (RD) between treatment and control groups when emulating a target trial of angiotensin converting enzyme inhibitor (ACEI) versus angiotensin receptor blocker (ARB) on the risk of primary cardiovascular events (CVD) in people with HIV (PWH) with an intention-to-treat (ITT) and per-protocol (PP) analysis.

We then tested for an interaction effect by race, which was not statistically significant for both PP (p=0.71) and ITT analysis (p=0.36), and fitted models stratified by race. ACEIs significantly reduced CVD risk for PWH in PP analysis, with similar effect for Black PWH (0.79 [0.55-1.14]) and White PWH (0.64 [0.45-0.90]). The observed effect was not statistically significant in the ITT analysis (Black PWH: 1.01 [0.85-1.20] ; White PWH: 0.84 [0.71-1.00]).

### ARBs vs. no antihypertensive treatment

The ITT analysis of ARB versus no treatment indicated a slight reduction in the relative hazard of CVD (0.87 [0.65-1.16]). The PP analysis of ARB versus no treatment showed a significant reduction for the relative hazard (0.37 [0.18-0.76]) of CVD.

The interaction between race and treatment was not statistically significant (p=0.22). For the stratified ITT analysis, ARB initiation was associated with a CVD hazard of 0.59 [0.33-1.06] in White PWH and 0.97 [0.63-1.49] in Black PWH. 5-year RDs can be found in Table 3.

### ACEIs vs. ARBs

The emulated target trials comparing the initiation of ARB versus ACEI showed no statistically significant difference in CVD risk. To avoid confusion in direct comparisons, all reported HR are defined the ratios of CVD hazard in the ARB initiators group over the hazard in ACEI initiators group. The ITT HR was 1.13 [0.81-1.57]. The PP analysis showed a non-statistically significant lower risk for ARB initiators (0.54 [0.25-1.18]). 5-year RDs can be found in Table 3.

The product term between treatment and race as part of an ITT analysis was not statistically significant (p=0.54). The estimated ITT effect of ACEI versus ARB spanned the null for both White PWH (0.84 [0.53-1.32]) and for Black PWH (0.92 [0.57-1.50]).

### Sensitivity analysis excluding the largest cohort

The analysis excluding the largest cohort yielded consistent results with the main analysis. Compared with no antihypertensive treatment, ACEIs and ARBs were similarly effective in all PWHs in preventing CVD events (ACEI ITT: 0.76 [0.65-0.88]; ACEI PP: 0.75 [0.56-1.00]; ARB ITT: 0.92 [0.66-1.27]; ARB PP: 0.33 [0.14-0.77]). Stratified analyses by race also remained consistent with the primary findings (see Web Appendix Table A2, Figure A4). However, it reduced the overall sample size, particularly in the ARB treatment group. Detailed analysis is provided in Web Appendix.

## 4. Discussion

The head-to-head comparison of ACEIs and ARBs revealed similar effectiveness for reducing CVD risk among PWH, consistent with findings among people without HIV (7,8,11). When compared to a third group of hypertensive PWH without treatment, both drugs significantly reduced the risk of CVD.

Our findings also addressed ongoing concerns about potential differences in the effectiveness across racial groups. Specifically, our results did not suggest a significant difference of these medications among Black compared to White PWH. While, on average, the initiation of ARB or ACEI appeared to be more protective for White PWH compared to Black PWH in both PP (for ACEI) and ITT (for both ACEI and ARB) analysis, observed differences were not significant and should be interpreted carefully.

This analysis contributes to answering the body of concerns raised in prior studies regarding the suitability of ACEIs and ARBs as first-line antihypertensive options for Black patients (17,32,33). Such questions are especially relevant given that Black individuals comprised 70% of new HIV infections in the United States in 2022 (34). Therefore, future research should explore the relative impact of these drugs compared to other antihypertensives within Black populations and compare the balance of benefits and harms of these drugs. Additionally future research could look into more detail in the impact of ARB for Black patients.

However, these findings should be interpreted with caution. A primary limitation is the lack of blood pressure measurements in our analysis, which prevents us from directly assessing the physiological impact of ARBs and ACEIs. Additionally, we did not collect detailed information on medication dosage, raising the possibility that Black PWH may have received lower or less effective doses of ACEIs or ARBs. This could have contributed to the observed differences in treatment effectiveness. The fractures within the U.S. healthcare system present further challenges. Individuals might receive care from multiple providers, often across different healthcare networks and settings. This fragmentation increases the risk of incomplete documentation and underreporting of prescribed medications, which in turn limits our ability to accurately identify the true initiation of antihypertensive therapy. Furthermore, inconsistencies in record-keeping can mean that the exact start and stop dates of medications are not reliably captured in the dataset. It is also possible that our data did not fully encompass all health interventions. Some individuals may have received non-pharmacological management for hypertension, such as lifestyle counseling, or may have intermittently used antihypertensive drugs outside of the ACEI or ARB classes. Such interventions would not be consistently recorded in our dataset, potentially leading to misclassification and dilution of the observed effects. Furthermore, information on drug use prior to NA-ACCORD enrollment was limited, which may affect accurate identification of primary antihypertensive therapy. Lastly, reported CVD diagnoses were not centrally validated in NA-ACCORD, except for MI in a subset of cohorts, which may have led to under- or overestimation of CVD events.

Collectively, these limitations complicate the identification and interpretation of potential differences observed between groups. Therefore, future research should further investigate the comparative effectiveness and benefit-harm balance of prescribing ACEIs and ARBs versus other antihypertensive agents in Black PWH, ideally considering dosage, adherence, and socio-economic factors, to better inform treatment guidelines for this disproportionately affected population.

The reliability of our results also relies on adherence to the target RCT protocol (35). Although we aimed to closely replicate this explicit protocol, certain key elements of RCTs could not be fully emulated due to inherent limitations of observational data. For instance, treatment allocation in our study was not randomized; as a result, our conclusions stand on the assumption of conditional exchangeability, addressed through adjustment for all known and measured confounders.

Due to a lack of detailed information regarding reasons for treatment switches, we classified any deviation from the initial treatment assignment (ACEI only, ARB only or no-antihypertensive treatment) as non-adherence. The observed differences between ITT and PP effect estimates were consistent with dilution of the ITT effect due to non-adherence; that is, some individuals assigned to the treatment group in our hypothetical trial with an ITT analysis stopped taking the treatment shortly after initiation, attenuating the observed effect. While some may argue that treatment changes associated with serious adverse events should not be treated as protocol deviations and censored, we do not expect this to strongly impact our analysis, as both ACEIs and ARBs have well-established safety profiles, and serious adverse effects are relatively uncommon with these medications (12,36).

Finally, one might argue that our analysis did not explicitly account for deaths (non-CVD related) as competing risks when estimating the effect of exposure on CVD outcomes. However, in our cohorts, such competing events were very rare. In the ITT analysis, there were only 291 non-CVD deaths over 320’059 person-months in the group assigned to ACEI, 16 events non-CVD deaths over 11’627 person-months in the ARB group and 1,298 non-CVD deaths over 513’815 person-months in the no-treatment group. Given these very low event rates, the potential bias from not using a dedicated competing risks model, such as the Fine-Gray approach, is likely negligible in this context.

To assess the robustness of our results and evaluate the influence of cohort composition, we repeated all analyses after excluding the largest cohort. Although this reduced the sample size and limited the precision of direct treatment comparisons, treatment versus no-treatment effects and patterns across racial groups remained largely unchanged, reinforcing the credibility and generalizability of our findings, and robustness to both residual confounding and differences in cohort composition.

Despite these comprehensive analyses, further studies are needed to address the limitations of our work. Such research could include a thorough evaluation of potential adverse effects and incorporate additional clinical variables for adjustment, as well as additional outcomes such as congestive heart failure (CHF), which was not available in our dataset.

In conclusion, our real-world study indicates that ACEIs and ARBs are both effective in preventing CVD events among individuals with HIV and hypertension. In head-to-head comparisons, both drug classes were similarly protective in the overall population and in race-stratified analyses.

### NOVELTY AND RELEVANCE

#### What is New?

We used a causal inference framework to compare ACEIs and ARBs for prevention of CVD in people living with HIV and hypertension, and estimate subgroup-specific causal effects among Black and White people.

#### What is relevant?

This study provides estimates on comparative effectiveness of two of the main anti-hypertensive drug classes for primary CVD prevention in this large observational cohort of adults receiving HIV care in the U.S. and Canada, and estimated per-protocol and intention-to-treat hazard ratios and 5-years difference in cumulative risk of cardiovascular events for different treatment strategies for hypertension, overall and stratified by race.

#### Clinical/Pathophysiological Implications?

Both ACEIs and ARBs effectively reduce cardiovascular risk in people with HIV and hypertension, supporting their use for the management of hypertension. Stratified analysis revealed no-statistically significant differences is risk reduction for both drugs in White PWH compared to Black PWH.

## Acknowledgements

We are grateful to the Swiss National Science Foundation and NA-ACCORD funders for their support in funding this study. We thank Aimee Freeman, NA-ACCORD Project Manager and IeDEA Administrative Coordinator, for her crucial support in data curation and access, as well as facilitating NA-ACCORD steering committee reviews.

## Ethics approval

The human subjects’ activities of the NA-ACCORD has been approved by the Johns Hopkins School of Medicine institutional review board (NA_00002683), as well as each participating NA-ACCORD cohort was granted ethics approval by their respective local institutional review boards, and the NA-ACCORD was approved by the Johns Hopkins School of Medicine institutional review board.

## Data availability

The complete data for this study cannot be publicly shared due to legal and ethical restrictions. Access to NA-ACCORD data is possible with approval of a research concept sheet directly from the NA-ACCORD owners.

